# Knowledge Base and Emerging Frontiers in Post-Tuberculosis Lung Disease Research (2015–2025): A Bibliometric Analysis Based on the Web of Science

**DOI:** 10.64898/2026.07.01.26357080

**Authors:** Tingting Zhu, Yinping Feng, Yunkai Dai, Jiali Yu, Zunjing Zhang, Zhongda Liu

**Author notes:** Corresponding authors (ZL); (ZZ). These authors contributed equally to this work (Tingting Zhu, Yinping Feng). These authors also contributed equally to this work (Zunjing Zhang, Zhongda Liu).

## Abstract

**Background:** Post-tuberculosis lung disease (PTLD) constitutes a substantial global public health burden, yet its overall research landscape has not been systematically quantified. This bibliometric study characterizes global research trends and the knowledge architecture of PTLD publications issued between 2015 and 2025.

**Methods:** We retrieved PTLD-related literature from the Web of Science Core Collection using search terms including “post-tuberculosis lung disease” and “tuberculosis sequelae”. Only English original articles were retained, yielding a final dataset of 353 publications. We adopted VOSviewer, CiteSpace and the Bibliometrix R package to visualize annual publication outputs, cross-country and institutional collaborations, author co-authorship networks, keyword co-occurrence patterns, and citation burst dynamics.

**Results:** Annual publications rose from 13 in 2015 to 67 in 2025, with 71.4% of all papers published from 2021 to 2025. The five most productive nations were the United States (72 papers), India (61), China (51), the United Kingdom (49), and South Africa (38). The US and UK occupied core hub positions in international collaboration networks. Leading institutions included Stellenbosch University (20 articles), the University of Cape Town (14), and the University of Liverpool/Liverpool School of Tropical Medicine (10). Keyword co-occurrence clustering identified eight thematic groups, with dominant hotspots covering post-tuberculosis bronchiectasis, pulmonary function impairment, chronic pulmonary aspergillosis (CPA), and quality of life. The keywords with the strongest recent citation bursts were “post-tuberculosis lung disease” (strength = 3.79), “prevalence” (3.70), and “quality of life” (2.51). Research frontiers extending to 2025 center on standardized PTLD conceptual framing, pulmonary rehabilitation, and long-term clinical outcome assessment.

**Conclusions:** PTLD research has shifted from merely describing structural lung damage toward standardized disease definitions, functional pulmonary testing, complication management, and patient quality-of-life improvement. Future research priorities should include prospective multicenter cohort studies, individualized pulmonary rehabilitation programs, and host-directed therapies to mitigate the global burden of chronic PTLD.

## Introduction

The WHO Global Tuberculosis Report 2025 estimated 10.7 million incident tuberculosis (TB) cases and 1.23 million TB-associated deaths worldwide in 2024, with approximately 155 million TB survivors currently living globally^[1]^. While anti-TB treatment coverage has reached record highs, accumulating evidence confirms that lung injury induced by “Mycobacterium tuberculosis” can persist or even progress after microbiological cure^[2–4]^.

In 2019, the First International Post-Tuberculosis Pulmonary Health Symposium formally defined PTLD as persistent chronic respiratory abnormalities partially or fully attributable to prior TB infection, regardless of symptomatic presentation^[5]^. Epidemiological surveys show that 34%-74% of TB survivors retain residual pulmonary dysfunction, and radiologically confirmed bronchiectasis affects 44%-50% of cured patients^[6–7]^. Such structural and functional lung lesions drastically elevate long-term mortality: PTLD patients carry a 3-6-fold higher mortality risk than the general population, especially within the first year following treatment completion^[6]^.

In 2019, TB accounted for 122 million global disability-adjusted life years (DALYs), nearly 47% (58 million) of which stemmed from post-TB sequelae rather than active TB infection^[8]^. Traditional burden assessments have long prioritized active TB, systematically underestimating the public health weight of persistent lung damage after bacteriological cure^[9–10]^. Although PTLD research has gained growing attention in recent years, existing studies remain fragmented: most focus narrowly on epidemiology and clinical characterization, lacking systematic integration of pathological mechanisms and disease subphenotypes^[11–12]^. Consensus is absent across critical domains, including immune-mediated lung injury pathways, precise disease subtyping, and standardized long-term management algorithms^[13–14]^.

Bibliometrics enables quantitative evaluation of large-scale scientific outputs to objectively map core contributors, research hotspots, and thematic evolution, delivering actionable evidence for disciplinary development and disease control strategies. In this study, we extracted PTLD-related publications from the Web of Science Core Collection spanning 2015 to 2025 and performed systematic analyses via VOSviewer, CiteSpace and Bibliometrix. By constructing co-occurrence matrices for countries, institutions, authors and keywords, we identified high-frequency keyword clusters and tracked the temporal evolution of research frontiers. This work aims to summarize the global PTLD research landscape, unpack cross-national and cross-institutional collaboration disparities, and generate evidence-based recommendations for basic translational research and optimized public health resource allocation ^[15]^.

## Materials and Methods

### Data Sources and Search Strategy

All data were extracted from the Web of Science Core Collection (WoSCC). To guarantee search completeness and reproducibility, we constructed a Topic (TS) search query incorporating terms covering PTLD and its key clinical phenotypes, with the following syntax: TS=(((“post-tuberculosis” OR “post tuberculosis” OR posttuberculosis) NEAR/5 (lung OR pulmonary OR respiratory OR airway OR bronchial) OR “post-tuberculosis lung disease” OR “post tuberculosis lung disease” OR ((tuberculosis OR “pulmonary tuberculosis”) NEAR/3 (sequelae OR sequela OR bronchiectasis OR fibrosis OR “airflow obstruction” OR “airway obstruction” OR “lung impairment” OR “pulmonary impairment” OR “lung function” OR spirometry OR “forced expiratory volume” OR “forced vital capacity”))).

The initial search retrieved 844 records. After filtering document type to “Article”, 547 entries remained. We manually screened titles and abstracts to exclude irrelevant studies, conference abstracts and duplicate entries. Further constraints limited the dataset to publications issued between 2015 and 2025 and written in English, leaving 353 articles for downstream bibliometric analysis. All searches and data exports were completed within a unified time window to avoid bias from real-time database updates, and raw data were standardized before analysis.

### Data Analysis and Visualization Methods

After finalizing the article dataset, multiple bibliometric tools were deployed to dissect the knowledge architecture and developmental trajectories of PTLD research. Microsoft Excel 2021 was used for descriptive statistics of annual publication trends, which delineated the field’s growth phases and cumulative output.

Extended bibliometric analyses were executed via the bibliometrix 4.1.3 package in R (version 4.3.1). This module assessed journal distribution, author and institutional collaboration networks, national research productivity, and keyword co-occurrence patterns to systematically map research hotspots and thematic shifts. Knowledge visualization maps were generated using VOSviewer 1.6.20 and CiteSpace 6.2.R6. Specifically, VOSviewer constructed co-occurrence networks for authors, institutions, countries and keywords, producing density and temporal overlay visualizations. CiteSpace detected keyword citation bursts and reference co-citation networks, delineating the foundational knowledge base and evolving research frontiers of PTLD.

## Results

### Annual Growth Trend of Publications

Fig. 1 illustrates the annual publication output and cumulative publication volume in the field of PTLD from 2015 to 2025. The number of annual publications increased from 13 in 2015 to 20 in 2020. In 2021, annual publications rose to 43, twice the number recorded in the previous year. Between 2022 and 2023, annual publications remained stable at 42–44 articles, before reaching 58 in 2024 and 67 in 2025. The cumulative number of publications grew from 13 in 2015 to 101 by 2020, and then to 353 by the end of 2025. In total, 71.4% of all publications (252 out of 353) were produced between 2021 and 2025.

**Fig. 1.**
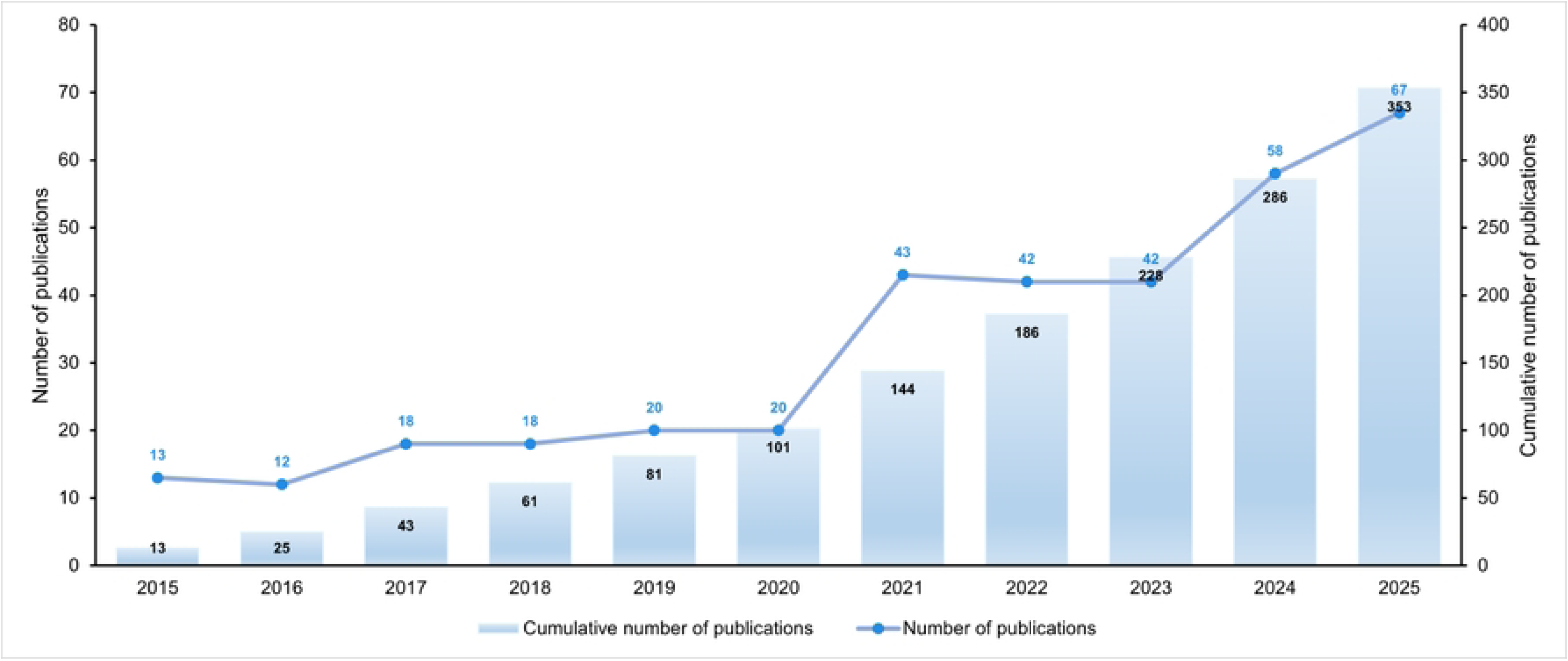
Annual and cumulative publication trends in PTLD research from 2015 to 2025. The bar chart shows the number of original articles published per year; the line graph indicates the cumulative number of publications. Data were retrieved from the Web of Science Core Collection. Abbreviation: PTLD, post-tuberculosis lung disease.

### National Productivity and Cross-Regional Collaboration Networks

PTLD research exhibits a multi-center global distribution stratified by publication volume, total citations, and collaborative linkage strength. The United States, India, China, the United Kingdom, and South Africa constitute the top five productive nations (Table 1). The United States leads with 72 articles, followed by India (61), China (51), the United Kingdom (49), and South Africa (38). Secondary contributors include South Korea (30), Brazil (24), Turkey (16), France (15), and Germany (13).

**Table 1.** Top 10 most productive countries in PTLD research (2015–2025)

| Rank | Country | Publications<br>(NP) | Total<br>Citations<br>(TC) | TLS | Avg.<br>Citations |
| --- | --- | --- | --- | --- | --- |
| 1 | USA | 72 | 1918 | 146 | 26.6389 |
| 2 | India | 61 | 722 | 26 | 11.8361 |
| 3 | China | 51 | 575 | 12 | 11.2745 |
| 4 | United<br>Kingdom | 49 | 1464 | 139 | 29.8776 |
| 5 | South Africa | 38 | 1099 | 105 | 28.9211 |
| 6 | South Korea | 30 | 412 | 13 | 13.7333 |
| 7 | Brazil | 24 | 302 | 25 | 12.5833 |
| 8 | Turkey | 16 | 40 | 0 | 2.5 |
| 9 | France | 15 | 449 | 49 | 29.9333 |
| 10 | Germany | 13 | 341 | 62 | 26.2308 |

From an impact perspective, the United States recorded the highest total citation count (1918) alongside a Total Link Strength (TLS) value of 146. The United Kingdom ranked second in total citations (1464, with a TLS value of 139), while South Africa recorded a relatively high average citation count of 28.92 per article. France and Germany also exhibited relatively high per-article citation values, with average citations of 29.93 and 26.23, respectively. In contrast, India and China, despite large publication output, displayed substantially lower average citation metrics (11.84 and 11.27). Turkey’s 16 articles accumulated only 40 total citations with no cross-border collaborative links (a TLS value of 0).

The national collaboration network (Fig. 2A) places the United States at the core, maintaining robust partnerships with the United Kingdom, South Africa, Germany, France, and Switzerland. The United Kingdom forms a secondary central hub, establishing tight cooperative ties with India, the Netherlands, Italy, and South Africa. China and South Korea form a distinct East Asian collaborative subnetwork, whereas India sustains extensive cross-border links with the United Kingdom, the Netherlands, Spain, and Singapore. South Africa exhibits high overall connectivity across the global network.

**Fig. 2.**
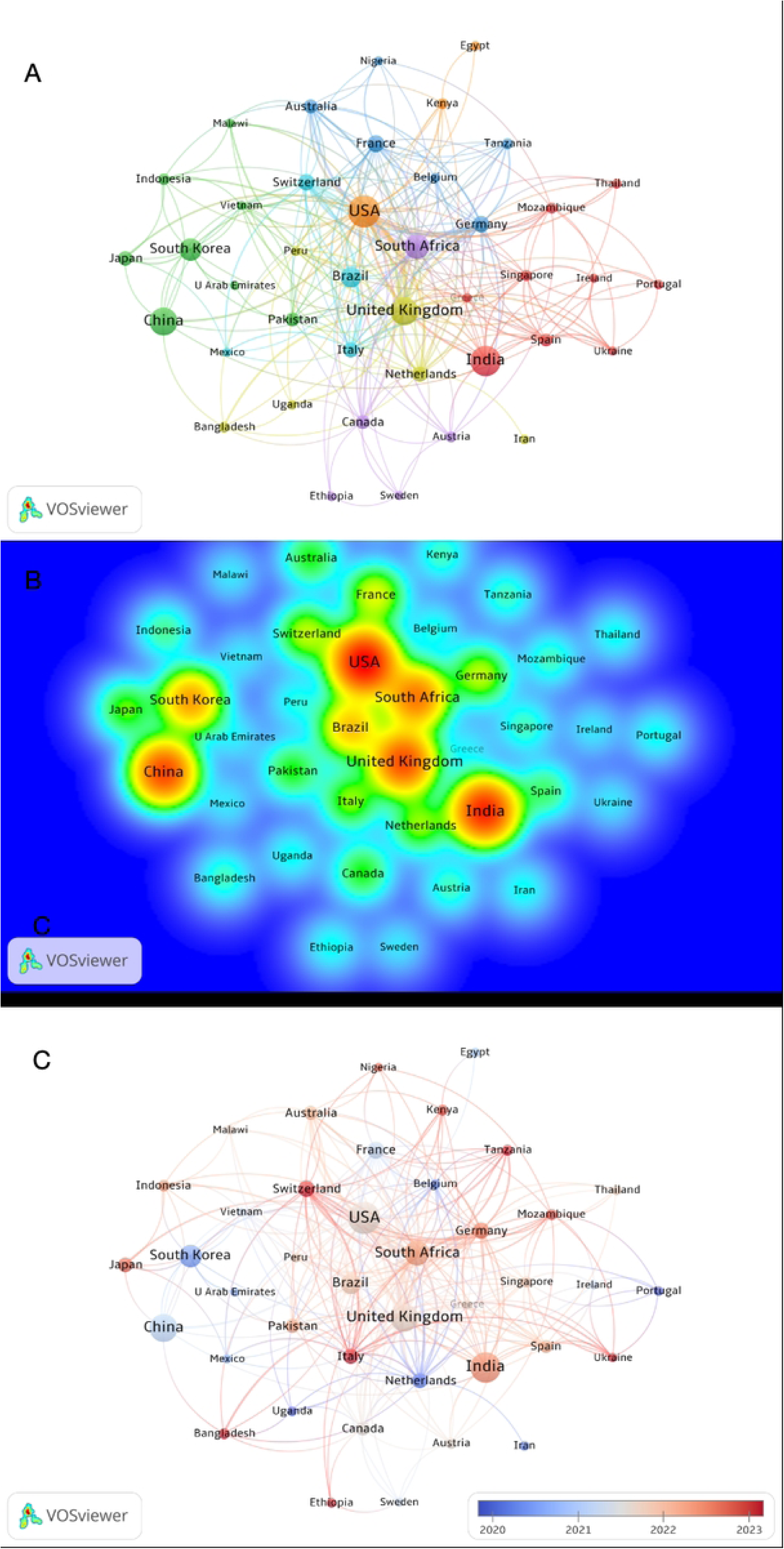
National collaboration network in PTLD research (2015–2025). (A) Network visualization of cooperative relationships among countries. Node size represents the number of publications, and edge thickness indicates the strength of collaboration. (B) Density visualization highlighting research hotspots; warmer colors (e.g., red) indicate higher density of collaboration and publication activity. (C) Temporal overlay visualization, where node colors transition from blue (earlier average publication year) to red (later average publication year), illustrating the temporal evolution of research activity across countries.

The national density heatmap (Fig. 2B) reveals concentrated research hotspots centered on the United States, India, China, the United Kingdom, and South Africa. The United States presents the highest collaborative density, with secondary high-intensity clusters distributed around the United Kingdom and South Africa. Additional hotspots are observed in France, Germany and South Korea, while peripheral nations including Portugal, Ukraine, Iran, Sweden, and Egypt occupy marginal positions within the network.

Temporal overlay visualization (Fig. 2C) differentiates early-stage contributors (cool blue nodes: China, South Korea, the Netherlands, Belgium, France) from nations with active research output in recent years (warm red nodes: India, Germany, Switzerland, Italy, Kenya, Tanzania, Mozambique, Thailand).

### Institutional Distribution and Collaboration Networks

Based on publication volume, total citations, and collaboration network data, institutional distribution in PTLD research reveals clear regional clustering, with core strengths concentrated in South Africa, the United Kingdom, the United States, and selected East Asian institutions (Table 2). Stellenbosch University ranked first with 20 publications, followed by the University of Cape Town (14 articles).

**Table 2.**
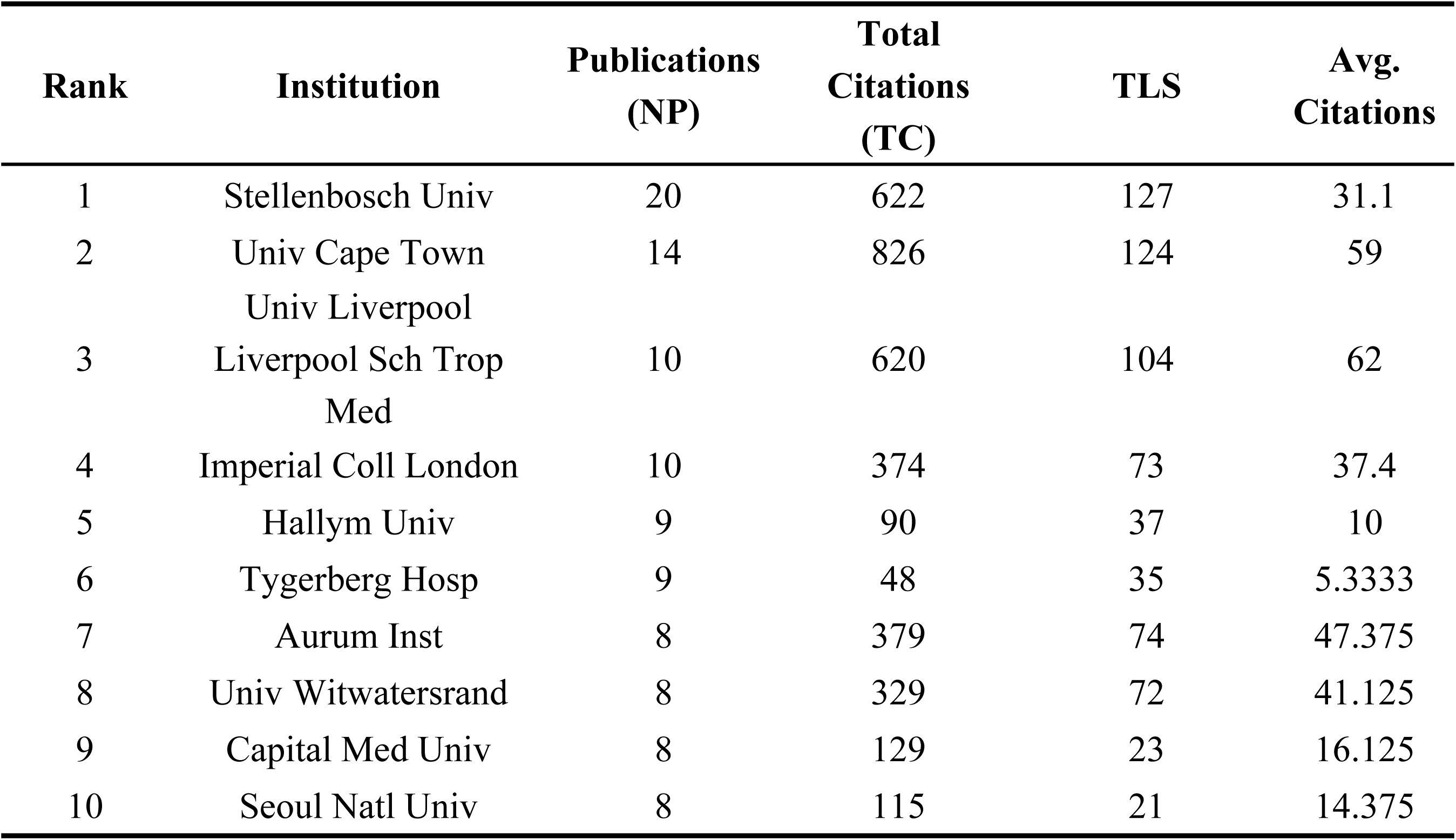
Top 10 most productive institutions in PTLD research (2015–2025)

The University of Liverpool/Liverpool School of Tropical Medicine and Imperial College London each contributed 10 articles (tied for third). Hallym University and Tygerberg Hospital each published 9 articles, while the Aurum Institute, the University of the Witwatersrand, Capital Medical University, and Seoul National University each produced 8 articles.

In terms of academic impact, the University of Cape Town had the highest total citations (826), with an average of 59.00 per article. The University of Liverpool/Liverpool School of Tropical Medicine achieved 620 total citations and the highest average among the top 10 (62.00). Stellenbosch University had 622 total citations and a TLS of 127. The Aurum Institute and the University of the Witwatersrand received 379 and 329 citations, with averages of 47.38 and 41.13, respectively. Asian institutions such as Capital Medical University, Seoul National University, and Hallym University exhibited lower citation metrics.

The collaboration network (Fig. 3A) revealed a core structure centered on South African institutions, closely linked with research groups in the United Kingdom and the United States. Stellenbosch University, the University of Cape Town, the University of the Witwatersrand, the Aurum Institute, and Tygerberg Hospital form a densely connected cluster, with Stellenbosch and Cape Town occupying central positions. UK institutions, including Imperial College London, the University of Liverpool/Liverpool School of Tropical Medicine, and the London School of Hygiene & Tropical Medicine, maintain strong ties with their South African counterparts.

**Fig. 3.**
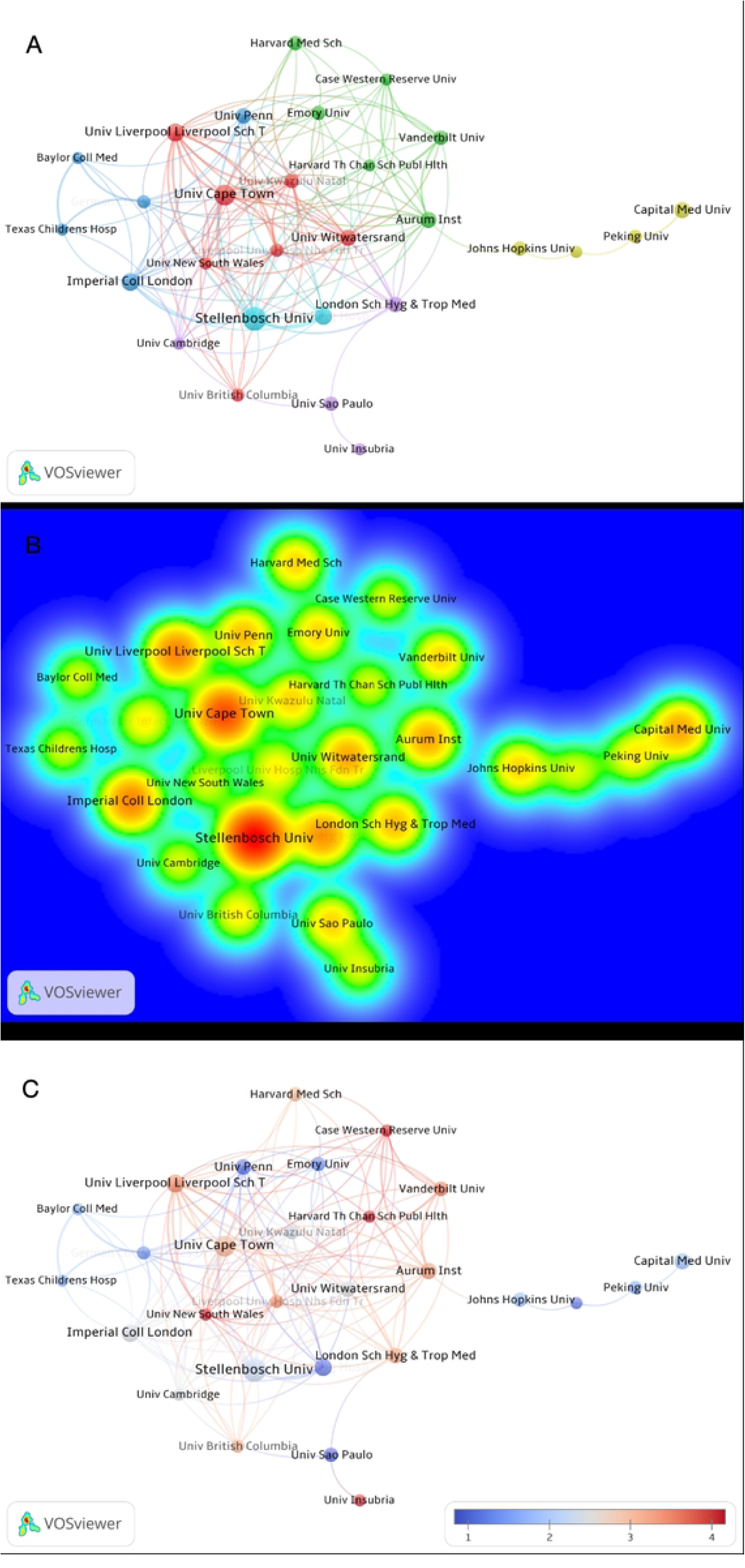
Institutional collaboration network in PTLD research (2015–2025). (A) Network visualization of cooperative relationships among institutions. Node size represents the number of publications, and edge thickness indicates the strength of collaboration (Total Link Strength, TLS). (B) Density visualization highlighting research hotspots; warmer colors (e.g., red) indicate higher density of collaboration and publication activity. (C) Temporal overlay visualization, where node colors transition from blue (earlier average publication year) to red (later average publication year), illustrating the temporal evolution of research activity across institutions.

Beyond the South Africa-UK core, US institutions such as Harvard Medical School, Emory University, Case Western Reserve University, Vanderbilt University, and Johns Hopkins University constitute another major collaborative hub, with multi-directional links to both South African and UK institutions. Chinese institutions, represented by Capital Medical University and Peking University, form a smaller but distinct sub-network, connecting to the main network through Johns Hopkins University.

The institutional density heatmap (Fig. 3B) identified research hotspots concentrated around Stellenbosch University, the University of Cape Town, the University of Liverpool/Liverpool School of Tropical Medicine, Imperial College London, the University of the Witwatersrand, the Aurum Institute, and the London School of Hygiene & Tropical Medicine. Stellenbosch and Cape Town displayed the highest intensity. Notably, Johns Hopkins University, Peking University, and Capital Medical University formed a secondary hotspot on the right side of the network.

The temporal overlay (Fig. 3C) demonstrated variation in node colors. Most core institutions presented intermediate colors (green), while institutions such as the University of New South Wales, Case Western Reserve University, and the University of Insubria exhibited warmer colors.

### Journal Distribution

Based on publication volume, total citation frequency, and journal co-occurrence network data, knowledge platforms for PTLD research are concentrated in respiratory medicine, tuberculosis, infectious disease, and general medical journals (Table 3).

**Table 3.** Top 10 most productive journals in PTLD research (2015–2025)

| Rank | Journal | Publications (NP) | Total Citations (TC) | TLS | Avg. Citations |
| --- | --- | --- | --- | --- | --- |
| 1 | BMC Pulmonary Medicine | 12 | 119 | 28 | 9.9167 |
| 2 | Cureus Journal Of Medical Science | 11 | 25 | 16 | 2.2727 |
| 3 | International Journal Of Chronic Obstructive Pulmonary Disease | 10 | 179 | 38 | 17.9 |
| 4 | Mycoses | 10 | 64 | 11 | 6.4 |
| 5 | Journal Of Thoracic Disease | 8 | 41 | 9 | 5.125 |
| 6 | PLOS One | 7 | 279 | 24 | 39.8571 |
| 7 | International Journal Of Tuberculosis And Lung Disease | 6 | 382 | 75 | 63.6667 |
| 8 | Journal Of Fungi | 6 | 111 | 11 | 18.5 |
| 9 | Journal Of Clinical And Diagnostic Research | 6 | 4 | 7 | 0.6667 |
| 10 | Thorax | 5 | 235 | 49 | 47 |

In terms of publication volume, BMC Pulmonary Medicine ranked first with 12 articles, followed by Cureus Journal of Medical Science (11 articles). The International Journal of Chronic Obstructive Pulmonary Disease and Mycoses each published 10 articles (tied for third). The Journal of Thoracic Disease published 8 articles, PLOS One published 7, while the International Journal of Tuberculosis and Lung Disease, the Journal of Fungi, and the Journal of Clinical and Diagnostic Research each published 6 articles, and Thorax published 5.

From an academic impact perspective, the International Journal of Tuberculosis and Lung Disease had a total citation count of 382, a TLS of 75, and an average of 63.67. Thorax had a total citation count of 235 and an average of 47.00. PLOS One had a total citation count of 279 and an average of 39.86. BMC Pulmonary Medicine had a total citation count of 119 and an average of 9.92. The International Journal of Chronic Obstructive Pulmonary Disease had a total citation count of 179 and an average of 17.90.

The journal co-occurrence network map (Fig. 4A) demonstrates that journals such as BMC Pulmonary Medicine, the International Journal of Tuberculosis and Lung Disease, the Journal of Thoracic Disease, PLOS One, and Mycoses feature larger nodes and more connections, positioned in the core area of the network. Specifically, the International Journal of Tuberculosis and Lung Disease is closely linked with journals related to respiratory diseases, public health, and infectious diseases. BMC Pulmonary Medicine and the Journal of Thoracic Disease are more connected to respiratory specialty and general clinical journals. Journals such as Mycoses, the Journal of Fungi, Infection and Drug Resistance, and Clinical Infectious Diseases form an infectious-disease-related subgroup on the right side of the network.

**Fig. 4.**
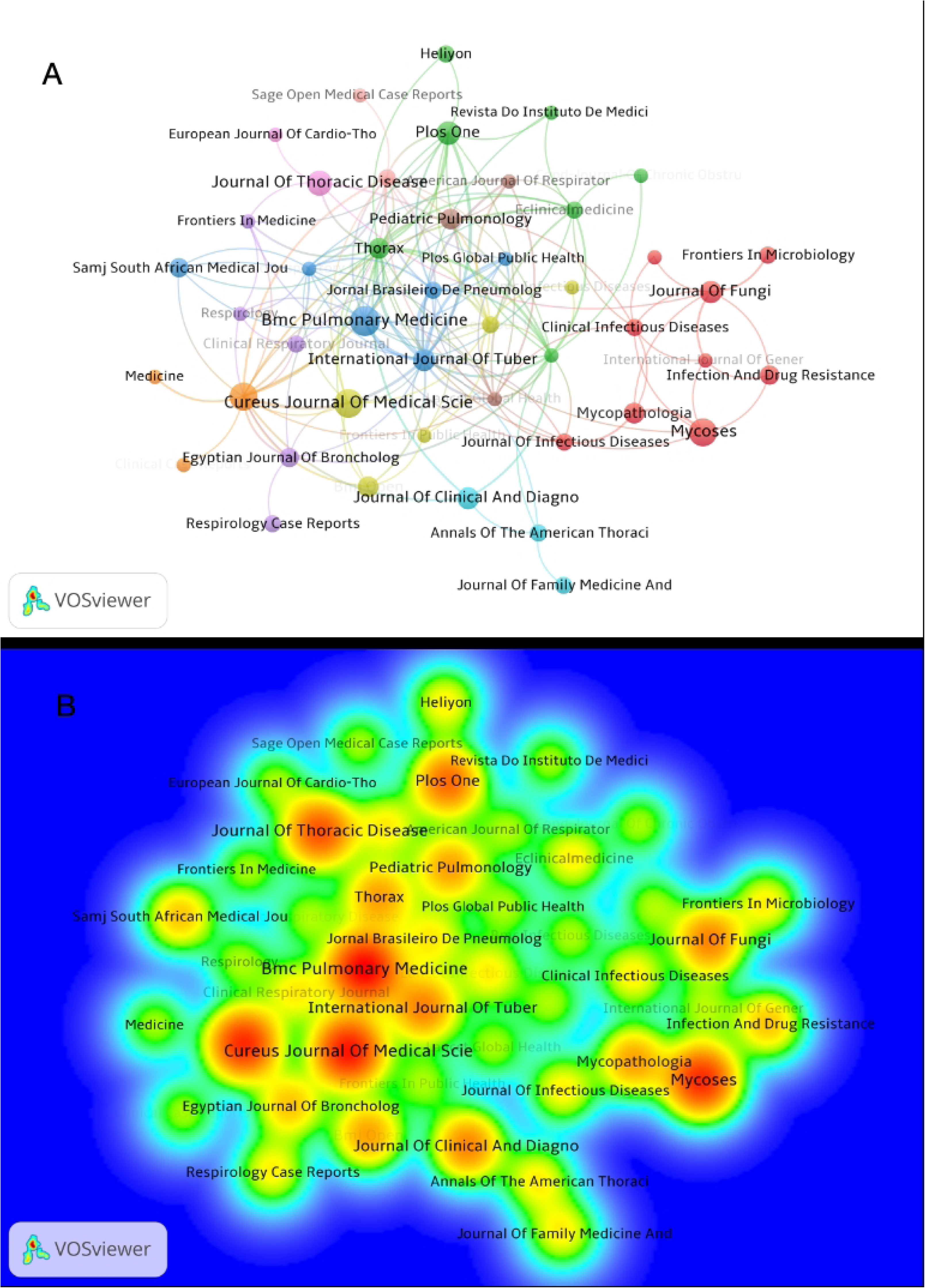
Journal co-occurrence network in PTLD research (2015–2025). (A) Network visualization of journal co-occurrence based on citation or co-citation relationships. Node size reflects the number of publications or total link strength, and edge thickness indicates the strength of co-occurrence links. (B) Density visualization showing the concentration of research themes; warmer colors (e.g., red) indicate higher density of journal co-occurrence, representing core journals or major research clusters.

Three distinct thematic journal clusters can be identified. The first category primarily focuses on respiratory diseases and pulmonary function impairment assessment (e.g., BMC Pulmonary Medicine, Thorax, the Journal of Thoracic Disease). The second category emphasizes tuberculosis control and public health management (e.g., the International Journal of Tuberculosis and Lung Disease, PLOS Global Public Health). The third category leans toward infectious disease and mycology research (e.g., Mycoses, the Journal of Fungi, Journal of Infectious Diseases). Comprehensive journals such as Cureus Journal of Medical Science and the Journal of Clinical and Diagnostic Research also occupy peripheral positions in the network.

The journal density heatmap (Fig. 4B) illustrates that research hotspots are concentrated around Cureus Journal of Medical Science, the International Journal of Tuberculosis and Lung Disease, Mycoses, the Journal of Thoracic Disease, BMC Pulmonary Medicine, and PLOS One. Among these, the regions corresponding to the International Journal of Tuberculosis and Lung Disease, Mycoses, and Cureus Journal of Medical Science exhibit warmer colors. Secondary hotspot zones form around journals such as Thorax, Pediatric Pulmonology, Clinical Infectious Diseases, and the Journal of Fungi.

### Author Collaboration Network and Core Research Groups

Based on author publication volume, total citation frequency, and author collaboration networks, PTLD research has formed several author collaboration groups. The overall landscape is characterized by multi-center parallel development, with no single dominant author team (Table 4).

**Table 4.** Top 10 most productive authors in PTLD research (2015–2025)

| Rank | Author | Publications<br>(NP) | Total<br>Citations<br>(TC) | TLS | Avg.<br>Citations |
| --- | --- | --- | --- | --- | --- |
| 1 | Agarwal, Ritesh | 9 | 66 | 86 | 7.3333 |
| 2 | Dhooria, Sahajal | 9 | 66 | 86 | 7.3333 |
| 3 | Muthu, Valliappan | 9 | 66 | 86 | 7.3333 |
| 4 | Sehgal, Inderpaul<br>Singh | 9 | 66 | 86 | 7.3333 |
| 5 | Prasad, Kuruswamy<br>Thurai | 8 | 56 | 82 | 7 |
| 6 | Chakrabarti, Arunaloke | 7 | 50 | 75 | 7.1429 |
| 7 | Aggarwal, Ashutosh<br>Nath | 7 | 51 | 71 | 7.2857 |
| 8 | Garg, Mandeep | 7 | 50 | 69 | 7.1429 |
| 9 | Wallis, Robert S. | 6 | 199 | 73 | 33.1667 |
| 10 | Rudramurthy,<br>Shivaprakash M. | 6 | 33 | 64 | 5.5 |

In terms of publication volume, Agarwal Ritesh, Dhooria Sahajal, Muthu Valliappan, and Sehgal Inderpaul Singh each published 9 articles. Prasad Kuruswamy Thurai published 8 articles. Chakrabarti Arunaloke, Aggarwal Ashutosh Nath, and Garg Mandeep each published 7 articles, while Wallis Robert S. and Rudramurthy Shivaprakash M. each published 6 articles.

From an academic impact perspective, Wallis Robert S. recorded a total citation frequency of 199, with an average of 33.17. Agarwal Ritesh, Dhooria Sahajal, Muthu Valliappan, and Sehgal Inderpaul Singh each recorded total citation frequencies of 66, with averages of 7.33. Prasad Kuruswamy Thurai obtained 56 citations and a TLS of 82. The TLS values for Chakrabarti Arunaloke, Aggarwal Ashutosh Nath, and Garg Mandeep were 75, 71, and 69, respectively.

The author collaboration network (Fig. 5A) reveals three core teams. The first core team lies in the lower-left region of the network, centered around Agarwal Ritesh, Dhooria Sahajal, Muthu Valliappan, Garg Mandeep, Aggarwal Ashutosh Nath, and Prasad Kuruswamy Thurai. The second team occupies the upper-middle part of the network, with Wallis Robert S. as the core, connecting with Churchyard Gavin, Bisson Gregory P., Bakuli Abhishek, Charalambous Salome, and Maenetje Pholo. The third collaboration group is situated on the right side of the network, centered around Allwood Brian, linking Baines Nicola, Goussard Pierre, Hatherill Mark, Griffith-Richards Stephanie, and Andronikou Savvas. In the upper-left area of the network, a smaller independent collaboration cluster exists, comprising Keir Holly R., Long Merete B., Horton Katie L., Perea Lidia, and Mcdonnell Melissa J.

**Fig. 5.**
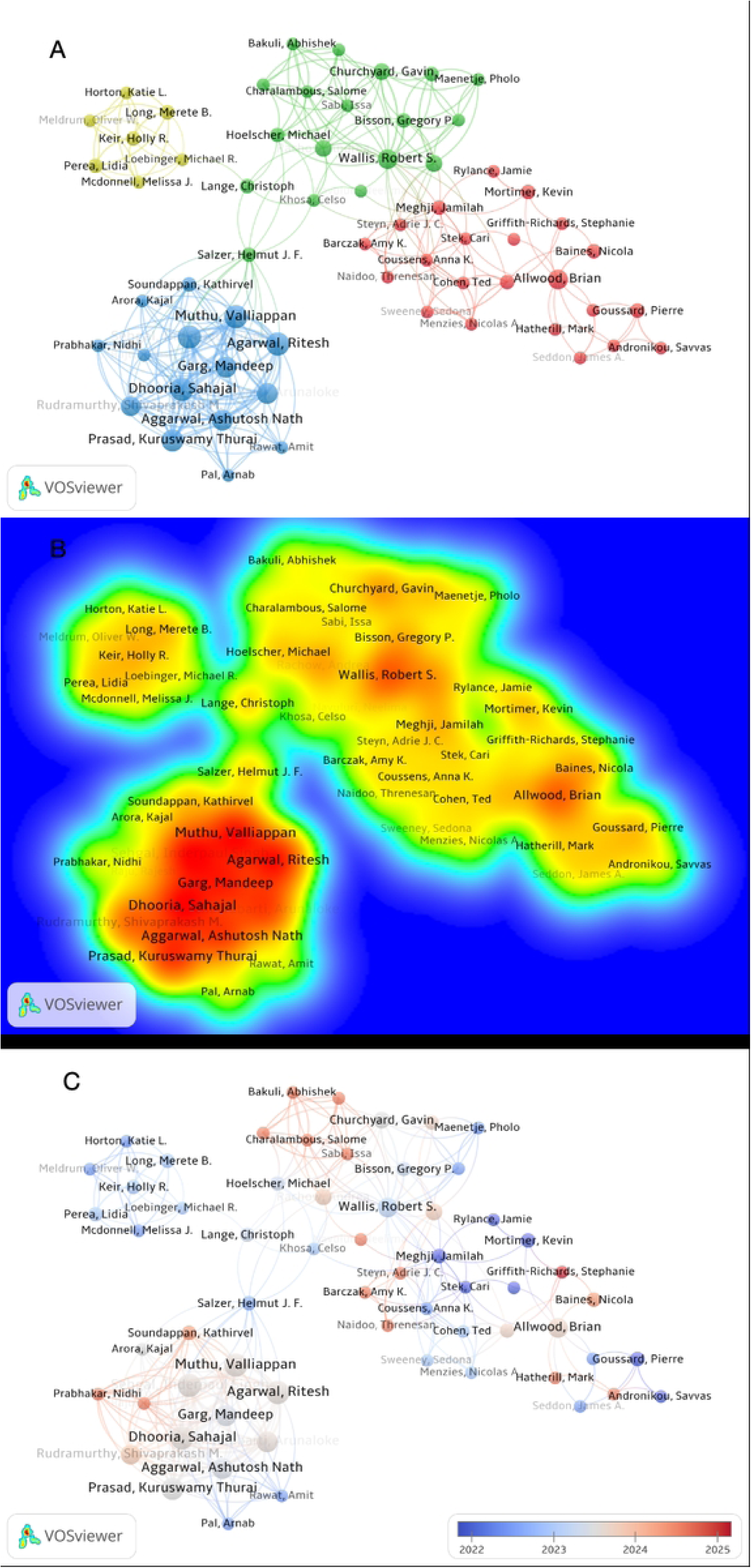
Author collaboration network in PTLD research (2015–2025). (A) Network visualization of co-authorship relationships among authors. Node size represents the number of publications, and edge thickness indicates the strength of collaboration. (B) Density overlay map highlighting research hotspots; warmer colors (e.g., red) indicate higher density of collaboration and publication activity. (C) Temporal overlay visualization, where node colors transition from blue (earlier average publication year) to red (later average publication year), illustrating the temporal evolution of research activity across authors.

Multiple sub-groups can be identified within the full author network, with interconnections established across different clusters. Wallis Robert S., Salzer Helmut J. F., and Allwood Brian act as bridging nodes connecting discrete research subgroups. Salzer Helmut J. F. links the lower-left author cluster to the central network region, whereas Allwood Brian maintains extensive internal connections within the right-side team and with peripheral authors.

The author density overlay map (Fig. 5B) illustrates research hotspots concentrated around Agarwal Ritesh, Dhooria Sahajal, Garg Mandeep, Aggarwal Ashutosh Nath, Prasad Kuruswamy Thurai, and Muthu Valliappan. Secondary hotspots emerge around Wallis Robert S. and Allwood Brian, and the lower-left author cluster presents the highest collaborative density.

The temporal overlay visualization (Fig. 5C) demonstrates clear differentiation of node color gradients. Nodes corresponding to Keir Holly R., Long Merete B., Horton Katie L., Perea Lidia, and Mcdonnell Melissa J. carry cooler blue tones. Nodes of Agarwal Ritesh, Dhooria Sahajal, Garg Mandeep, Aggarwal Ashutosh Nath, and Prasad Kuruswamy Thurai display warmer red hues. Within the right-side collaborative group, Griffith-Richards Stephanie and Baines Nicola also feature warmer node colors.

### Citation Analysis and Knowledge Base

Based on high-citation references, document co-citation networks, and citation burst detection, the knowledge base of PTLD research centers on imaging manifestations, pulmonary structural damage, pulmonary function abnormalities, disease burden assessment, and the conceptual construction of PTLD (Table 5).

**Table 5.** Top 10 most cited articles in PTLD research (2015–2025)

| Rank | Paper | DOI | Total Citations (TC) | TC per Year | Normalized TC |
| --- | --- | --- | --- | --- | --- |
| 1 | NACHIAPPAN AC, 2017, RADIOGRAPHICS | 10.1148/rg.2017160032 | 227 | 22.7 | 5.03 |
| 2 | PANDA A, 2017, DIAGN INTERV RADIOL | 10.5152/dir.2017.16454 | 225 | 22.5 | 4.99 |
| 3 | AMARAL AFS, 2015, EUR RESPIR J | 10.1183/13993003.02325-2014 | 167 | 13.92 | 3.13 |
| 4 | ALLWOOD BW, 2020, INT J TUBERC LUNG D | 10.5588/ijtld.20.0067 | 167 | 23.86 | 3.83 |
| 5 | MENZIES NA, 2021, LANCET GLOB HEALTH | 10.1016/S2214-109X(21)00367-3 | 158 | 26.33 | 10.96 |
| 6 | KWAN BW, 2015, ENVIRON MICROBIOL | 10.1111/1462-2920.12873 | 157 | 13.08 | 2.95 |
| 7 | SINGH R, 2018, PLOS ONE | 10.1371/journal.pone.0204155 | 140 | 15.56 | 5.92 |
| 8 | MEGHJI J, 2020, THORAX | 10.1136/thoraxjnl-2019-213808 | 138 | 19.71 | 3.17 |
| 9 | DODD PJ, 2021, LANCET INFECT DIS | 10.1016/S1473-3099(20)30919-1 | 108 | 18 | 7.49 |
| 10 | PORVAZNIK I, 2017, ADV EXP MED BIOL | 10.1007/5584_2016_45 | 98 | 9.8 | 2.17 |

In terms of highly cited publications, the article by Nachiappan AC (2017, Radiographics) records the highest total citations (227), followed by Panda A (2017, Diagnostic and Interventional Radiology) with 225 citations. The articles by Amaral AFS (2015, European Respiratory Journal) and Allwood BW (2020, International Journal of Tuberculosis and Lung Disease) each have 167 total citations. The article by Menzies NA (2021, Lancet Global Health) received 158 citations, with an annual citation rate of 26.33 and a normalized citation count of 10.96. The article by Dodd PJ (2021, Lancet Infectious Diseases) has 108 total citations and a normalized citation count of 7.49.

The document co-citation network (Fig. 6A) illustrates that publications by Allwood BW from 2020 and 2021 lie at the network’s center, featuring large nodes and extensive interconnections. Publications by Meghji J (2020), Menzies NA (2021), Migliori GB (2021), Nightingale R (2023), and Dodd PJ (2021) also occupy the core region. Earlier cited works include Denning DW (2016), van Kampen SC (2018), Ravimohan S (2018), Page ID (2019), Graham BL (2019), and Romanowski K (2019), and several of these nodes carry purple outer rings representing high betweenness centrality.

**Fig. 6.**
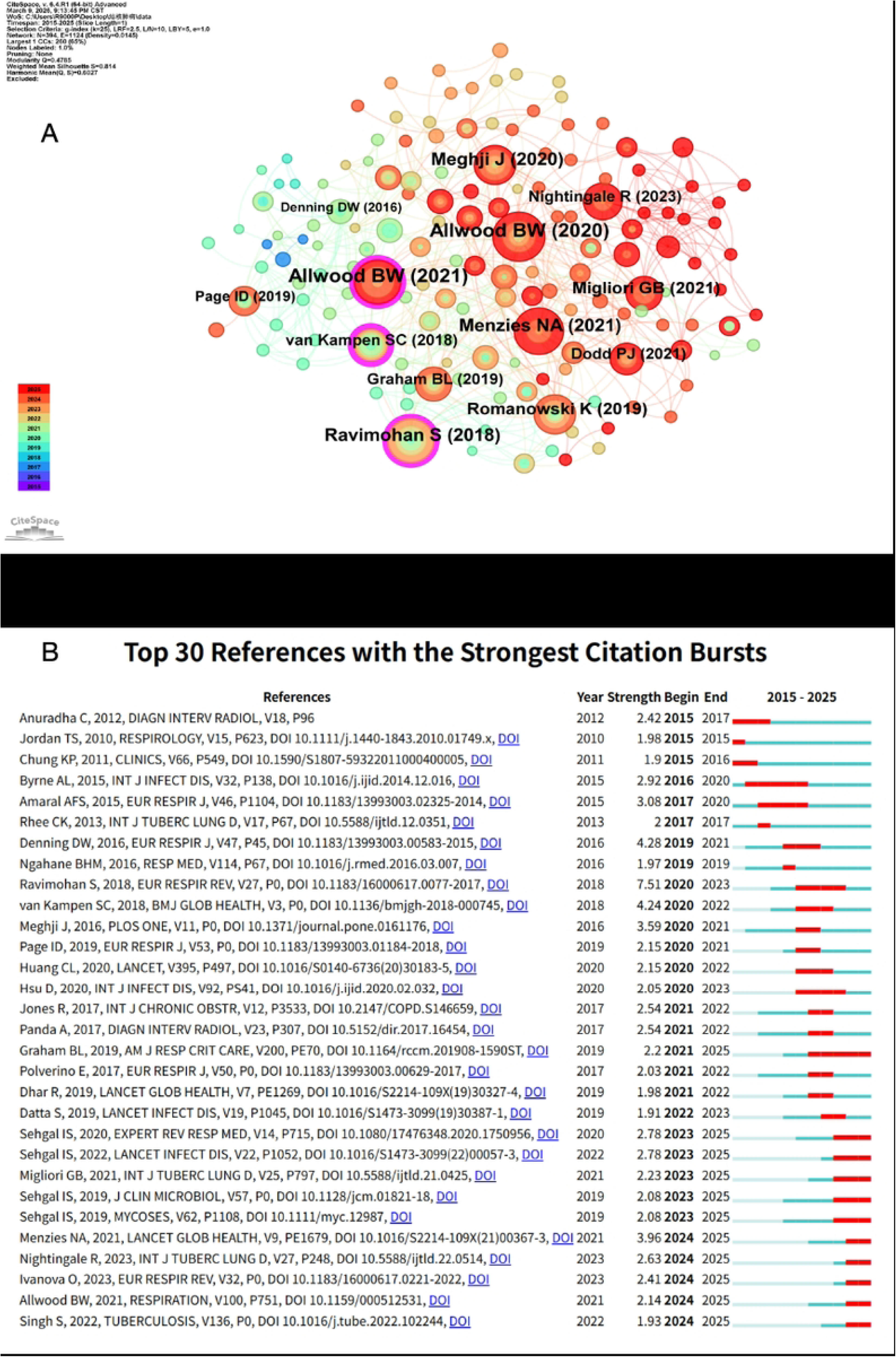
Co-citation network and citation burst analysis of references in PTLD research. (A) Document co-citation network. Nodes represent cited references, with node size proportional to total citation count. Nodes marked with purple outer rings indicate high betweenness centrality, acting as critical intellectual bridges linking distinct research clusters. (B) Citation burst visualization for references with the most prominent burst signals, where green line segments correspond to each reference’s publication lifespan, and red segments denote the timing, length and magnitude of citation bursts.

Citation burst analysis (Fig. 6B) identified 30 references with significant citation bursts across 2015-2025. Early burst publications active from 2015 to 2017 included Anuradha C (2012), Jordan TS (2010), and Chung KP (2011). Between 2016 and 2020, works such as Byrne AL (2015) and Amaral AFS (2015) exhibited prominent citation bursts. After 2020, the volume of burst references increased, accompanied by prolonged burst durations.

In terms of burst strength, the work by Ravimohan S (2018, European Respiratory Review) achieved the maximum strength value of 7.51, with a burst window spanning 2020-2023. Denning DW (2016, European Respiratory Journal) recorded a strength of 4.28 (2019–2021); van Kampen SC (2018, BMJ Global Health) scored 4.24 (2020–2022); Menzies NA (2021, Lancet Global Health) reached 3.96 (2024–2025); and Meghji J (2016, PLOS One) had a burst strength of 3.59 (2020–2021).

A large set of burst references retain active citation trends through 2025, including Graham BL (2019), Sehgal IS (2019, 2020, 2022), Migliori GB (2021), Menzies NA (2021), Nightingale R (2023), Ivanova O (2023), Allwood BW (2021), and Singh S (2022).

### Keyword Co-occurrence Analysis and Research Hotspots

Based on keyword frequency, network structure, and temporal overlay visualization results, PTLD research hotspots revolve around disease definition, structural lung damage, pulmonary function assessment, chronic complications, and rehabilitation management (Fig. 7A, Fig. 7B, Fig. 7C).

**Fig. 7.**
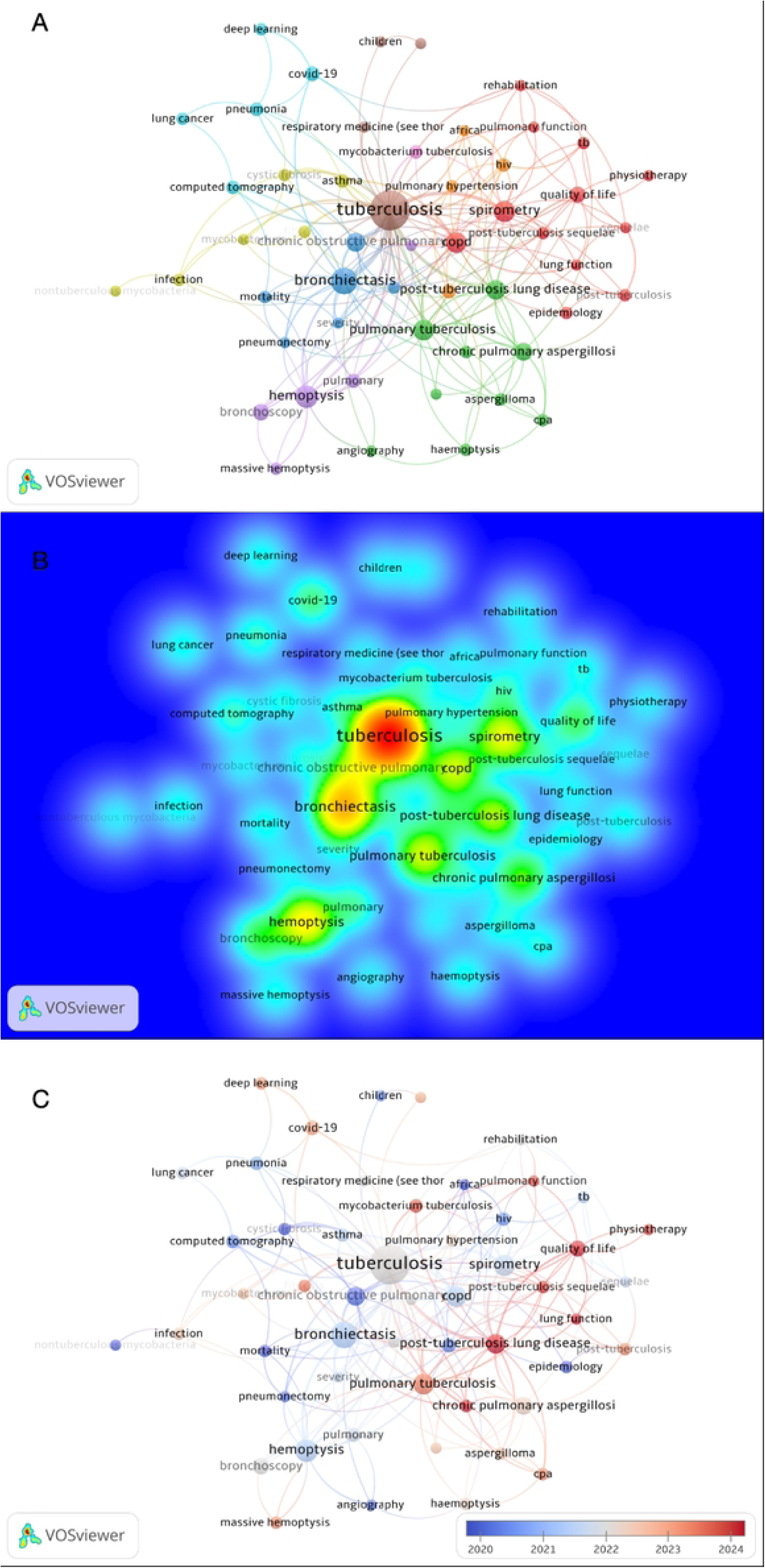
Keyword co-occurrence network in PTLD research (2015–2025). (A) Network visualization of keyword co-occurrence. Node size represents keyword frequency, and edge thickness indicates the strength of co-occurrence relationships. (B) Density overlay map highlighting research hotspots; warmer colors (e.g., red) indicate higher density of keyword co-occurrence. (C) Temporal overlay visualization, where node colors transition from blue (earlier average appearance year) to red (later average appearance year), illustrating the temporal evolution of research topics.

Among high-frequency keywords, “tuberculosis” appeared 89 times, with a TLS of 336. “Bronchiectasis” appeared 33 times, “hemoptysis” 22 times, “spirometry” 19 times, “COPD” 17 times, “pulmonary tuberculosis” 17 times, “post-tuberculosis lung disease” 15 times, “chronic obstructive pulmonary disease” 14 times, “chronic pulmonary aspergillosis” 12 times, and “bronchoscopy” 10 times. “Chronic obstructive pulmonary disease” had an average citation frequency of 17.00, and “chronic pulmonary aspergillosis” had an average citation frequency of 13.58.

The keyword co-occurrence network (Fig. 7A) reveals several distinct research clusters. The first cluster centers on “post-tuberculosis lung disease”, “spirometry”, “quality of life”, “pulmonary function”, “rehabilitation”, and “physiotherapy”. The second cluster is built around “pulmonary tuberculosis”, “chronic pulmonary aspergillosis”, “aspergilloma”, “CPA”, and “haemoptysis”. The third keyword group consists of “bronchiectasis”, “COPD”, “mortality”, “pneumonectomy”, and “severity”. A small fourth cluster takes “hemoptysis”, “bronchoscopy”, “massive hemoptysis”, and “angiography” as core nodes. Supplementary associated keywords include “computed tomography”, “pneumonia”, “COVID-19”, “deep learning”, and “lung cancer”.

In terms of network centrality, “tuberculosis” maintains extensive linkages with “bronchiectasis”, “spirometry”, “COPD”, “post-tuberculosis lung disease”, and “pulmonary tuberculosis”. “Bronchiectasis” is positioned in the lower-central area of the network, while “spirometry” and “post-tuberculosis lung disease” occupy the core zone on the network’s right side.

The keyword density overlay map (Fig. 7B) illustrates that research hotspots concentrate around “tuberculosis”, “bronchiectasis”, “spirometry”, “post-tuberculosis lung disease”, “pulmonary tuberculosis”, “hemoptysis”, and “COPD”. The regions corresponding to “tuberculosis” and “bronchiectasis” exhibit the warmest color gradients, and “spirometry” together with “post-tuberculosis lung disease” forms a secondary high-density hotspot.

Temporal overlay visualization (Fig. 7C) differentiates keywords by node color. Cool-toned blue nodes cover “bronchiectasis”, “hemoptysis”, “computed tomography”, “infection”, “mortality”, and “pneumonectomy”. Warm red nodes correspond to emerging research themes including “post-tuberculosis lung disease”, “post-tuberculosis sequelae”, “quality of life”, “pulmonary function”, “rehabilitation”, “physiotherapy”, “chronic pulmonary aspergillosis”, and “aspergilloma”. Recent frontier keywords such as “COVID-19” and “deep learning” also appear within the network.

### Keyword Clustering Structure and Thematic Evolution

Based on CiteSpace keyword clustering and temporal evolution analysis, the keyword network yielded a modularity Q value of 0.4785 and a weighted mean silhouette S value of 0.814 (Fig. 8A, Fig. 8B, Fig. 8C). Eight primary clusters were identified: #0 pulmonary tuberculosis, #1 surgery, #2 health status, #3 subacute invasive aspergillosis, #4 pulmonary rehabilitation, #5 cystic fibrosis, #6 post-tuberculosis lung disease, and #7 respiratory function tests.

**Fig. 8.**
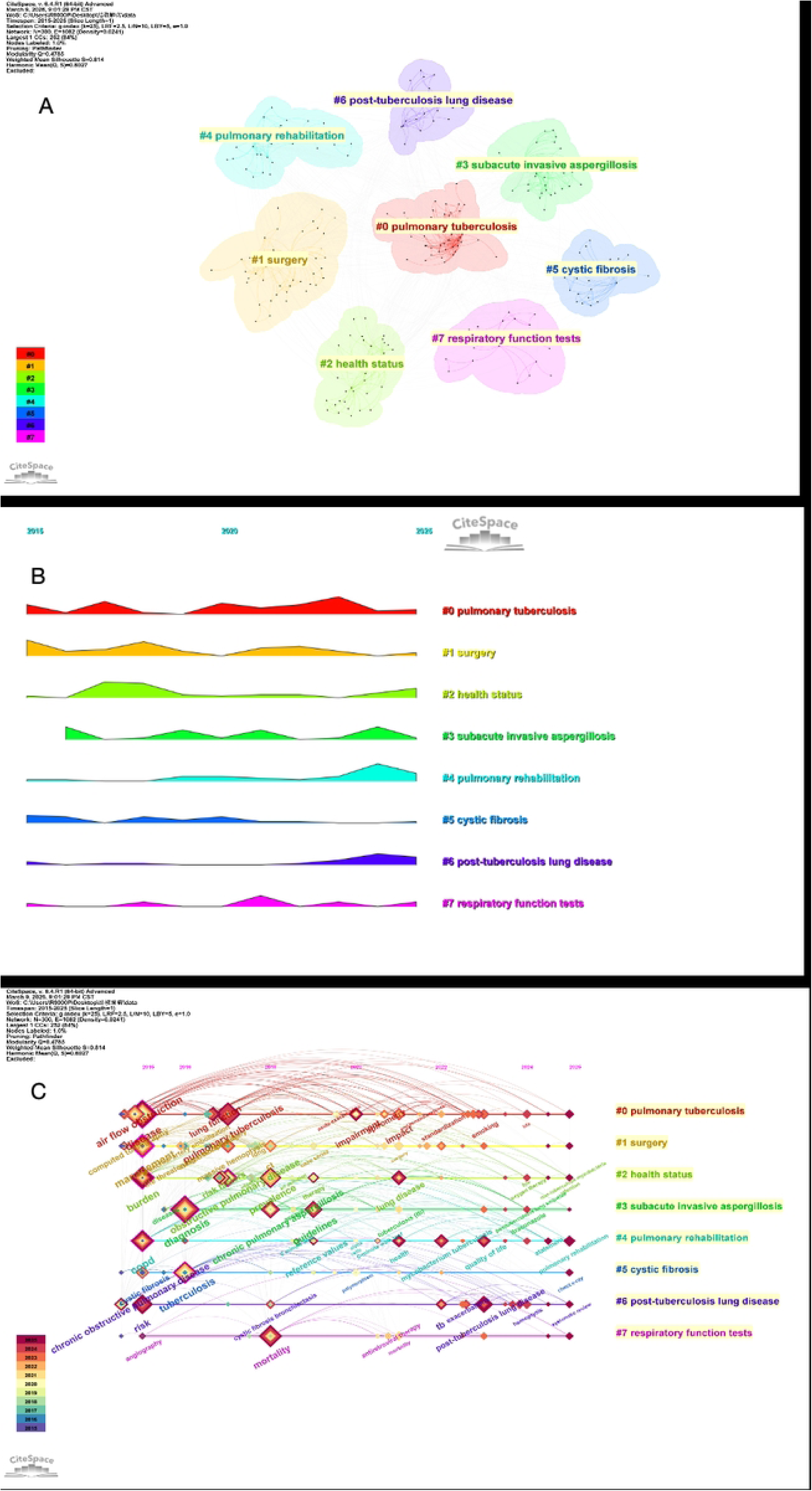
Keyword clustering structure and thematic evolution in PTLD research (2015–2025). (A) Keyword cluster visualization generated via CiteSpace. Nodes are partitioned into eight distinct clusters (#0–#7), each representing an independent core research theme. (B) Cluster activity mountain map reflecting temporal research popularity for each cluster across the study period; peaks indicate time intervals with intensive research output. (C) Keyword timeline visualization. Keywords belonging to identical clusters are arranged horizontally according to their average emergence year, visualizing long-term shifts and frontiers of PTLD research themes.

In terms of cluster distribution (Fig. 8A), #0 pulmonary tuberculosis sits at the network center and features the largest scale. This cluster covers related keywords including pulmonary function, airflow obstruction, functional impairment, and standardized assessment. The #1 surgery cluster centers on severe structural lung injury, hemoptysis, and surgical or interventional management strategies. The #2 health status cluster corresponds to disease burden, population morbidity, long-term prognosis, and persistent chronic health outcomes. The #3 subacute invasive aspergillosis cluster focuses on chronic and subacute fungal secondary infections after tuberculosis. The #6 post-tuberculosis lung disease cluster directly corresponds to the core research disease entity. The #4 pulmonary rehabilitation cluster incorporates rehabilitation interventions, exercise tolerance, quality-of-life improvement, and standardized clinical management protocols.

The keyword cluster mountain map (Fig. 8B) illustrates that #0 pulmonary tuberculosis sustains stable high research activity across the entire observation window. The #1 surgery cluster exhibits prominent activity in the early and middle phases of the timespan. The #2 health status cluster presents periodic activity fluctuations, with a clear revival in recent years. The #3 subacute invasive aspergillosis cluster displays multiple discrete activity peaks. The #4 pulmonary rehabilitation cluster reaches a distinct activity peak in the late period. The #6 post-tuberculosis lung disease cluster shows a marked upward trend in recent research output. The #7 respiratory function tests maintain steady continuous activity throughout the decade.

The keyword timeline graph (Fig. 8C) demonstrates distinct thematic shifts across three stages. From 2015 to 2018, research priorities centered on airflow obstruction, computed tomography imaging, clinical diagnosis, population disease burden, and mortality outcomes. Between 2018 and 2021, active keywords included chronic pulmonary aspergillosis, epidemiological prevalence, risk factors, quality of life, and normative reference values. From 2022 onward, novel nodes concentrated on post-tuberculosis lung disease, pulmonary rehabilitation, clinical practice guidelines, consensus statements, quality of life, long-term oxygen therapy, and systematic reviews appear on the right side of the timeline.

Notably, clusters #6 post-tuberculosis lung disease and #4 pulmonary rehabilitation display sustained extensible research momentum in the latter timeline segment. Cluster #3 subacute invasive aspergillosis maintains steady progressive development. By contrast, #5 cystic fibrosis and early diagnostic-related themes present weaker sustained research continuity in recent years.

### Thematic Trend Analysis and Frontier Shifts

Based on the thematic trend map (Fig. 9), early high-frequency research themes centered on cystic fibrosis, epidemiology, mortality, risk factors, chronic obstructive pulmonary disease, and computed tomography. Circa 2019, research priorities shifted toward pulmonary functional impairment and complex infectious or respiratory complications. Themes including HIV, tuberculosis, bronchiectasis, spirometry, and hemoptysis retained high research activity throughout 2020–2022.

**Fig. 9.**
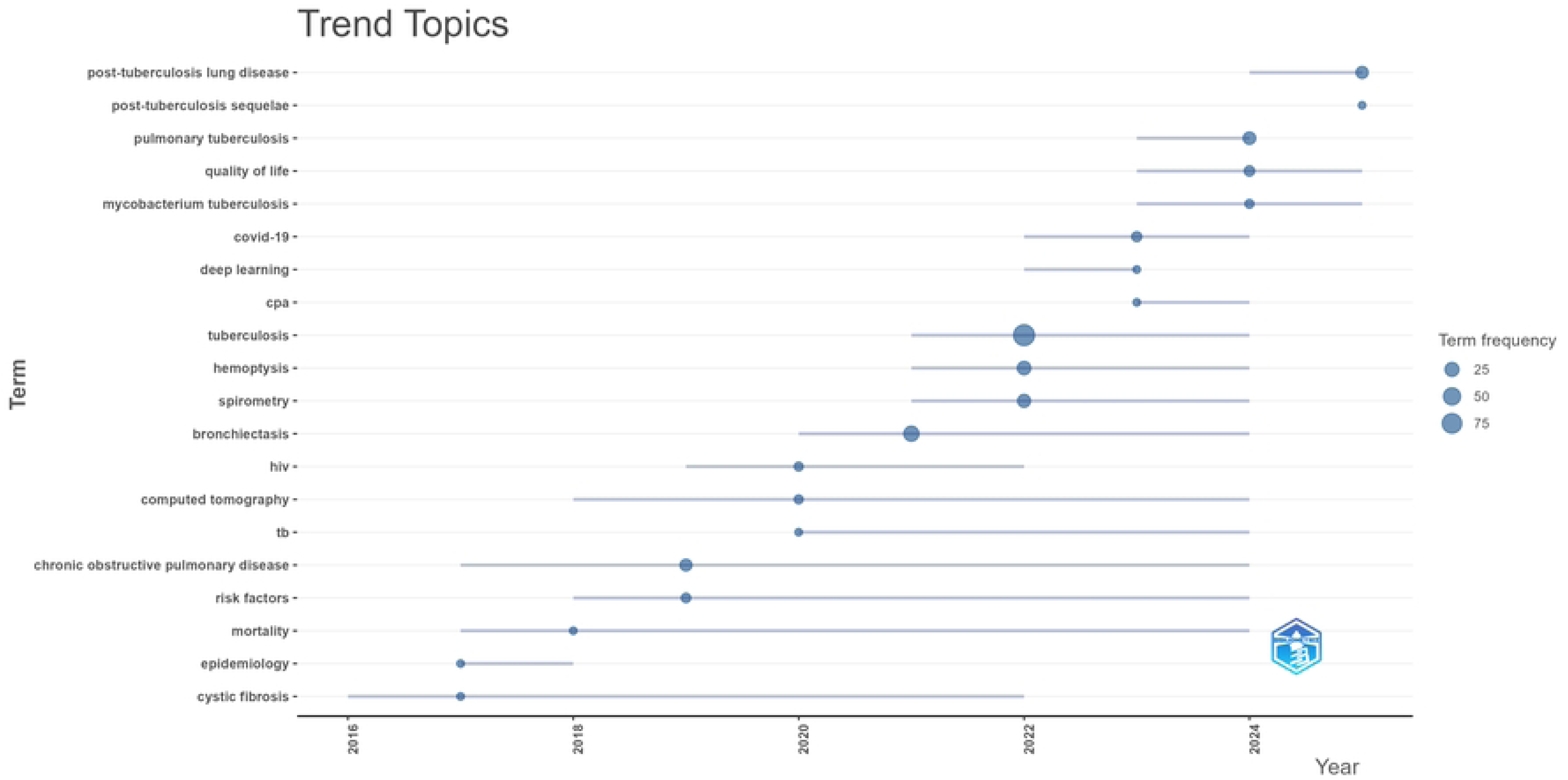
Thematic evolution trends in PTLD research (2015–2025). The horizontal axis denotes publication years. Each horizontal line corresponds to one distinct research theme, and circular markers record the year of theme emergence; larger circles signify higher keyword frequency. Vertical sorting of thematic tracks reflects sequential temporal shifts of PTLD research frontiers.

From 2022 onward, emerging active themes encompassed COVID-19, deep learning, CPA, Mycobacterium tuberculosis, quality of life, pulmonary tuberculosis, post-tuberculosis sequelae, and post-tuberculosis lung disease, with multiple topics sustaining research output through 2025. Of these thematic branches, post-tuberculosis lung disease occupies the rightmost segment of the timeline.

Pulmonary tuberculosis and Mycobacterium tuberculosis have maintained consistent research activity spanning 2023 to 2025. The CPA research theme gained prominence after 2023, while deep learning and COVID-19 represent recently emerging frontier directions.

In terms of sustained thematic lifespan, computed tomography, chronic obstructive pulmonary disease, risk factors, mortality, and bronchiectasis have remained active across a prolonged research window. By contrast, post-tuberculosis lung disease, post-tuberculosis sequelae, quality of life, and CPA emerged at a later stage yet rose rapidly in research popularity.

Regarding theme duration, computed tomography, chronic obstructive pulmonary disease, risk factors, mortality, and bronchiectasis persist over an extended period. Post-tuberculosis lung disease, post-tuberculosis sequelae, quality of life, and CPA appear later but ascend rapidly.

### Keyword Burst Analysis and Research Frontier Identification

Based on keyword burst analysis (Fig. 10), 30 keywords with prominent burst strength were detected across 2015-2025. From 2015 to 2019, “chronic obstructive pulmonary disease” recorded a burst strength of 2.29, “cystic fibrosis” 2.17, and “management” 1.82. During 2016–2018, “life-threatening hemoptysis” attained a burst strength of 3.15. The keyword “prevalence” showed a peak burst strength of 3.70 over 2018–2022, while “chronic pulmonary aspergillosis” displayed citation bursts spanning 2019–2021. Circa 2021, a series of new active burst keywords emerged, including “surgery”, “impact”, “lung function”, “function impairment”, and “morbidity”.

**Fig. 10.**
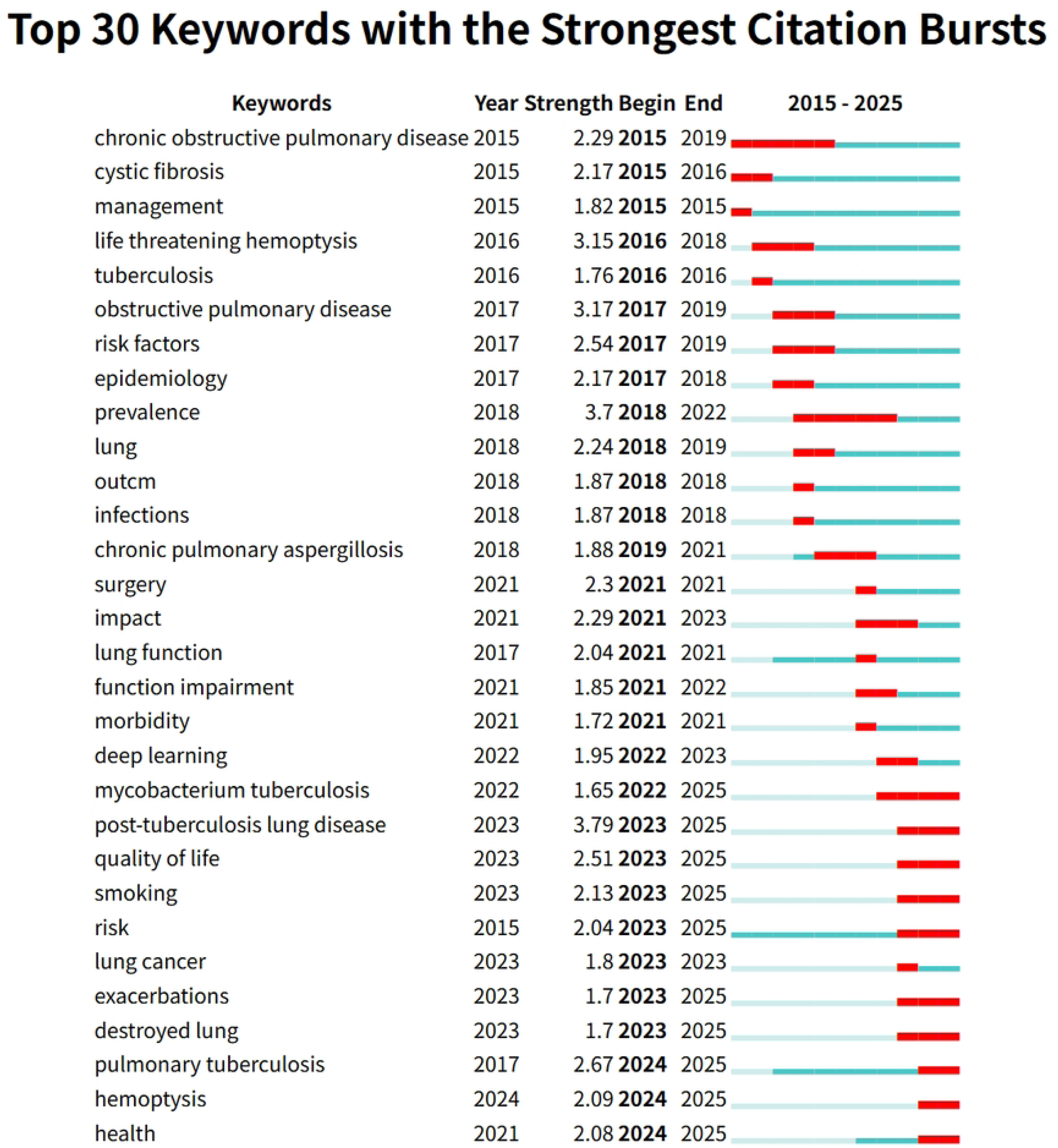
Top 30 keywords with the strongest citation bursts in PTLD research (2015–2025). Each row presents the keyword, its first occurrence year, burst strength, burst start year, and burst end year, paired with a horizontal bar timeline. Red bars mark periods of active keyword bursts, and blue bars denote non-burst periods.

During 2022–2025, “Mycobacterium tuberculosis” sustained continuous citation bursts through the end of the research window. “Post-tuberculosis lung disease” initiated a burst in 2023 with a strength value of 3.79, remaining active until 2025. “Quality of life”, “smoking”, and “risk” yielded burst strengths of 2.51, 2.13, and 2.04 respectively, all maintaining burst activity through 2025. Additional keywords retaining active bursts after 2023 covered “exacerbations”, “destroyed lung”, “pulmonary tuberculosis”, “hemoptysis”, and “health”.

When ranked by burst strength, “post-tuberculosis lung disease” took the lead, followed by “prevalence” (3.70), “obstructive pulmonary disease” (3.17), and “life-threatening hemoptysis” (3.15). The full set of frontier burst keywords persisting to 2025 comprises “post-tuberculosis lung disease”, “quality of life”, “smoking”, “risk”, “exacerbations”, “destroyed lung”, “pulmonary tuberculosis”, “hemoptysis”, and “health”.

## Discussion

### Paradigm Shift in PTLD Research

The most prominent temporal inflection point of PTLD research appeared in 2021, with 71.4% of all included articles published in the final five years of the study window. Such rapid research expansion temporally coincided with the First International Post-Tuberculosis Symposium held in 2019 and the subsequent unified definition and standardization of PTLD terminology, implying that consistent conceptual framing has greatly stimulated relevant research output^[16–17]^. More importantly, our keyword co-occurrence and cluster evolutionary analyses reveal a clear disciplinary paradigm shift: early research primarily focused on structural morphological characterization, dominated by keywords such as “bronchiectasis”, computed tomography (CT) and surgical intervention, where radiological evaluation of residual post-tuberculosis lesions was well-established. Intermediate-stage research centered on quantitative pulmonary functional evaluation, represented by spirometry, chronic obstructive pulmonary disease (COPD) and population prevalence surveys. Current research has gradually evolved toward holistic patient-centered clinical management, with core hotspots including the standardized PTLD disease framework, quality of life and pulmonary rehabilitation^[18–22]^. This developmental trajectory is consistent with the research evolution pattern of other chronic respiratory disorders, indicating that PTLD has transformed from a neglected secondary tuberculous sequela into an independent disease entity with standardized clinical intervention pathways^[23–24]^.

### Global Collaboration Landscape: From Productivity Disparity to the Know-Do Gap

Country-level and institutional bibliometric analyses demonstrate that the United States, the United Kingdom and South Africa act as core international knowledge hubs, while the global collaborative network also presents obvious imbalanced structural characteristics^[25–26]^. Although India and China rank among the top five nations in total publication volume, their average citation per article is substantially lower than that of South Africa and Western developed countries. This productivity-influence paradox proves that sheer publication quantity cannot directly translate into high global academic recognition. This phenomenon further reflects a widespread know-do gap within PTLD research: high-tuberculosis-burden middle-income countries can generate locally applicable clinical evidence, yet insufficient participation in transnational cooperative networks prevents such regional findings from being incorporated into global clinical guidelines and public health policy formulation^[27–28]^. South Africa provides a replicable development model, achieving both abundant research output and outstanding academic impact through long-term stable north-south cooperation with British and American research teams.

### Emerging Research Frontiers

Keyword burst detection further identifies multiple emerging research frontiers beyond the overall paradigm transformation of the field^[29]^. The standardized definition of “post-tuberculosis lung disease” exhibits the highest burst strength and forms an independent research cluster, marking a critical transition from scattered descriptions of isolated pulmonary residual lesions to an integrated, unified disease research framework^[16–17]^. Such conceptual consensus lays a necessary foundation for the development of PTLD-targeted therapeutic regimens, unified clinical evaluation criteria and public health policy recognition^[30]^. Sustained citation bursts of “destroyed lung” and “exacerbations” challenge the traditional static understanding of PTLD. Instead of stable, inactive late-stage sequelae, a large proportion of patients experience progressive functional deterioration and recurrent acute exacerbations throughout long-term follow-up^[31]^. Clinically, this evidence supports a COPD-style long-term management strategy for PTLD patients, including baseline spirometry-based clinical phenotyping, regular monitoring of exacerbation frequency and proactive preventive interventions, rather than a single routine follow-up after anti-tuberculosis treatment completion^[32]^. Pulmonary rehabilitation constitutes a large independent research cluster with a recent prominent activity peak, representing a research shift toward interventional functional recovery. Notably, our dataset contains the earliest published tele-rehabilitation clinical trials targeting PTLD populations^[33–34]^. However, most high-tuberculosis-burden regions lack specialized pulmonary rehabilitation institutions. Digital and community-based rehabilitation schemes hold transformative clinical potential, while critical research gaps remain regarding standardized exercise protocols, intervention duration and long-term adherence among PTLD patients, which deserve priority exploration^[35]^. Continuous research activity of “chronic pulmonary aspergillosis (CPA)” as an isolated cluster confirms that fungal complications are not rare niche manifestations but core components of the full PTLD clinical spectrum. Previous studies have clearly clarified the pathogenic mechanism: post-tuberculosis pulmonary cavities provide a favorable ecological niche for persistent Aspergillus colonization^[36–37]^. Nevertheless, our bibliometric results indicate routine CPA screening has not yet become standardized clinical practice for PTLD survivors. Co-occurring bursts of “hemoptysis” and “CPA” highlight an essential clinical diagnostic pathway: unexplained hemoptysis in cured tuberculosis patients requires systematic CPA screening, instead of being simply attributed to bronchiectasis.

### Relationship with Existing Literature

Our overall research conclusions support the unified PTLD conceptual framework proposed by Allwood et al., as well as the standardized clinical recommendations formulated by Migliori et al.^[16–17]^. The high centrality of these two landmark publications within the document co-citation network validates their foundational disciplinary status. Beyond reproducing existing consensus statements, the present study quantitatively visualizes the practical evolution of core PTLD concepts across the global research community. For instance, Migliori et al. proposed standardized pulmonary functional assessment standards, yet our keyword timeline analysis shows that spirometry and corresponding reference value research only gained widespread attention after 2021, reflecting an evident time lag between guideline release and large-scale clinical implementation. Furthermore, recently sustained frontier keywords including “quality of life” and “smoking” reflect a research shift toward multi-factor risk stratification. PTLD long-term outcomes are determined not only by initial tuberculosis lesion severity but also by modifiable risk factors such as tobacco exposure and socioeconomic status, consistent with findings from large population cohort studies^[18–19]^.

### Clinical and Policy Implications

This bibliometric analysis yields several actionable clinical and policy recommendations. For frontline clinicians, the growing focus on pulmonary function and patient-reported outcome measures (PROMs) suggests routine post-tuberculosis follow-up should integrate standardized spirometry and quality-of-life evaluation, rather than relying solely on chest radiography for lesion monitoring^[22,32]^. For clinical researchers, identified frontier directions including tele-rehabilitation, standardized CPA screening algorithms and long-term management of COPD-like recurrent exacerbations represent high-value targets for subsequent randomized controlled trials^[33,34,36]^. For public health policymakers, the unmet demand for standardized PTLD long-term care requires urgent investment from major global funding bodies such as the Global Fund to Fight AIDS, Tuberculosis and Malaria^[27]^. The inclusion of post-tuberculosis sequelae within the Global Burden of Disease (GBD) framework marked a landmark step to systematically quantify the global disease burden^[28]^. Even so, our bibliometric evidence reveals substantial barriers to translating such epidemiological evidence into standardized post-treatment care pathways within national tuberculosis control programs. Existing mathematical modeling studies confirm delayed anti-tuberculosis treatment exacerbates irreversible lung damage and all-cause mortality. Even under timely standard anti-tuberculosis therapy, disability-adjusted life years (DALYs) attributable to PTLD account for more than half of the total tuberculosis-related DALYs, emphasizing the critical need for early comprehensive intervention[29]. The worldwide clinical relevance of PTLD is further underscored by population estimates indicating approximately 155 million tuberculosis survivors were alive globally in 2020, 18% of whom completed anti-tuberculosis treatment within the preceding five years^[30]^.

### Limitations

Several methodological limitations of this bibliometric work must be acknowledged. First, literature retrieval restricted to the Web of Science Core Collection and English-language publications may introduce database and linguistic bias, potentially excluding regional non-English journals and clinical research conducted in high-burden low and middle-income countries. Second, bibliometric metrics cannot capture unpublished negative trial results, ongoing clinical studies and grey literature, which may skew observed research trends toward positive, publishable outcomes. Third, citation-based indicators are inherently time-dependent; newly published high-quality articles inevitably carry relatively low cumulative citation counts. We partially mitigate this limitation through normalized citation metrics and keyword burst detection algorithms. Fourth, although keyword clustering and burst analyses systematically map global research hotspots and frontiers, bibliometric methods cannot establish causal or mechanistic relationships. All pathophysiological interpretations within this discussion section are derived from cited original clinical research rather than direct outputs of the bibliometric analysis itself.

### Future Directions

Based on the visualized evolution of research frontiers, we propose four targeted directions for subsequent PTLD research. First, large-scale prospective multi-center cohort studies adopting unified PTLD diagnostic criteria are required to accurately quantify cross-regional disease burden across diverse tuberculosis-endemic populations^[16,29]^. Second, pragmatic randomized controlled trials of pulmonary rehabilitation—especially low-cost tele-rehabilitation schemes suitable for resource-limited settings—should be prioritized^[33–34]^. Third, implementation science research is needed to integrate routine CPA screening (Aspergillus IgG antibody testing and thoracic CT) into standardized post-tuberculosis follow-up algorithms^[36–37]^. Fourth, health economic research should evaluate the cost-effectiveness of diverse long-term monitoring models, given that optimal follow-up frequency and examination modalities remain undetermined^[27,30]^.

## Conclusion

In summary, this comprehensive bibliometric analysis demonstrates that PTLD research experienced rapid disciplinary maturation from 2015 to 2025. The research focus has gradually shifted from simple morphological documentation of post-tuberculosis lung damage toward a unified PTLD disease framework, standardized pulmonary functional assessment, targeted complication intervention and long-term quality-of-life optimization^[16,17,28]^. Looking beyond 2025, the most promising research frontiers cover pulmonary rehabilitation (particularly tele-rehabilitation), COPD-style chronic management of progressive lung dysfunction and recurrent exacerbations, and standardized screening protocols for fungal pulmonary complications^[33,34,36]^. Addressing these critical research priorities requires not only high-quality clinical trials but also sustained investment in transnational collaborative infrastructure, to narrow the persistent know-do gap between high-impact academic research institutions and high-tuberculosis-burden clinical populations worldwide^[27,30]^.

## Data Availability

All datasets analyzed in this study were retrieved from the Web of Science Core Collection database (https://www.webofscience.com). The complete search protocol, raw exported datasets, and analysis scripts can be obtained from the corresponding author upon reasonable request.

## Declarations

### Ethics approval and consent to participate

Not applicable. This study is a bibliometric analysis based on publicly available literature data retrieved from the Web of Science Core Collection database. No human subjects, animal experiments or individual medical records were involved, so ethical approval and informed consent were not required.

### Consent for publication

Not applicable. This manuscript contains no data from any individual person.

### Clinical trial number

Not applicable.

### Competing interests

The authors declare no competing interests.

### Authors’ contributions

Tingting Zhu and Yinping Feng contributed equally to this work and share first authorship. Zhongda Liu and Zunjing Zhang are co-corresponding authors. Conceptualization: Zhongda Liu, Zunjing Zhang; Data curation: Tingting Zhu, Yinping Feng; Formal analysis: Tingting Zhu, Yunkai Dai; Methodology: Jiali Yu, Yunkai Dai; Software: Yunkai Dai; Writing-original draft: Tingting Zhu, Yinping Feng; Writing-review & editing: Zhongda Liu, Zunjing Zhang. All authors read and approved the final manuscript.

## Acknowledgements

Not applicable.

## Notes

### Competing Interest Statement

The authors have declared no competing interest.

### Funding Statement

Yes

### Author Declarations

Not applicable. This is a bibliometric analysis based on publicly available data from the Web of Science Core Collection. No human participants, animal subjects, or personal data were involved, so no ethical approval was required.

